# Characteristics of three different chemiluminescence assays for testing for SARS-CoV-2 antibodies

**DOI:** 10.1101/2020.11.05.20225003

**Authors:** Myriam C. Weber, Martin Risch, Sarah L. Thiel, Kirsten Grossmann, Susanne Nigg, Nadia Wohlwend, Thomas Lung, Dorothea Hillmann, Michael Ritzler, Francesca Ferrara, Susanna Bigler, Konrad Egli, Thomas Bodmer, Mauro Imperiali, Yacir Salimi, Felix Fleisch, Alexia Cusini, Sonja Heer, Harald Renz, Matthias Paprotny, Philipp Kohler, Pietro Vernazza, Lorenz Risch, Christian R. Kahlert

## Abstract

Several tests based on chemiluminescence immunoassay techniques have become available to test for SARS-CoV-2 antibodies. There is currently insufficient data on serology assay performance beyond 35 days after symptoms onset. We aimed to evaluate SARS-CoV-2 antibody tests on three widely used platforms. A chemiluminescent microparticle immunoassay (CMIA; Abbott Diagnostics, USA), a luminescence immunoassay (LIA; Diasorin, Italy), and an electrochemiluminescence immunoassay (ECLIA; Roche Diagnostics, Switzerland) were investigated. In a multi-group study, sensitivity was assessed in a group of participants with confirmed SARS-CoV-2 (n=145), whereas specificity was determined in two groups of participants without evidence of COVID-19 (i.e. healthy blood donors, n=191, and healthcare workers, n=1002). Receiver operating characteristic (ROC) curves, multilevel likelihood ratios (LR), and positive (PPV) and negative (NPV) predictive values were characterized. Finally, analytical specificity was characterized in samples with evidence of Epstein–Barr virus (EBV) (n=9), cytomegalovirus (CMV) (n=7) and endemic common cold coronavirus infections (n=12) taken prior to the current SARS-CoV-2 pandemic. The diagnostic accuracy was comparable in all three assays (AUC 0.98). Using the manufacturers’ cut-offs, the sensitivities were 90%, 95% confidence interval,[84,94] (LIA), 93% [88,96] (CMIA), and 96% [91,98] (ECLIA). The specificities were 99.5% [98.9,99.8](CMIA) 99.7% [99.3,99,9] (LIA) and 99.9% [99.5,99.98] (ECLIA). The LR at half of the manufacturers’ cut-offs were 60 (CMIA), 82 (LIA), and 575 (ECLIA) for positive and 0.043 (CMIA) and 0.035 (LIA, ECLIA) for negative results. ECLIA had higher PPV at low pretest probabilities than CMIA and LIA. No interference with EBV or CMV infection was observed, whereas endemic coronavirus in some cases provided signals in LIA and/or CMIA. Although the diagnostic accuracy of the three investigated assays is comparable, their performance in low-prevalence settings is different. Introducing gray zones at half of the manufacturers’ cut-offs is suggested, especially for orthogonal testing approaches that use a second assay for confirmation.

## Introduction

COVID-19 is a recently emerging pandemic disease caused by infection with SARS-CoV-2 virus. Although there are considerable differences regarding its incidence, hospitalization rate, morbidity rate, and case fatality rate between different countries, disease control at the moment is uniformly achieved by strict prevention measures such as social distancing, wearing of face masks, hand-washing, contact tracing and testing, quarantine or isolation ^1^. Whereas the diagnosis of acute disease in the medical laboratory most importantly relies on RT-PCR testing, serological testing for antibodies specifically directed against viral proteins of SARS-CoV2 has increasingly come into the focus of public health authorities and medical institutions ^2-8^. The clinical presentation of COVID-19 has been shown to be heterogeneous in severity as well as in clinical signs and symptoms ^9^. A substantial proportion of patients have only minimal symptomatology or are even asymptomatic ^10^. Currently, there is no vaccination available, and causal therapies are very limited ^11^. After several countries showed success in confining the progression of the COVID-19 pandemic with drastic measures, there is a need to gradually cancel these measures to reinstitute possibly “normal” social and economic circumstances ^12,13^. To guide these actions to reverse socioeconomic lockdown, knowledge on the prevalence of COVID-19 is needed ^14^. Since a large proportion of patients with suspected COVID-19 infection could not be tested by RT-PCR in the acute phase, serological testing may gain increasing importance for retrospective clarification of clinical symptoms ^15^.

Serologic testing allows us to estimate the proportion of individuals already infected with COVID-19, either in the total population, in healthcare workplace settings or in general workplace settings ^16,17^. It facilitates contact tracing as well as surveillance and assists in the identification of individuals susceptible to COVID-19 infection ^2,18,19^. Furthermore, individuals having had contact with confirmed COVID-19 patients might be interested in determining whether they developed SARS-CoV-2 specific antibodies, if they did not yet have access to testing. Moreover, serological testing allows us to clarify clinical cases in which RT-PCR testing has been negative despite a high pretest probability for the presence of COVID-19 ^20,21^. Such cases of false negative RT-PCR have been reported, possibly because of improper collection techniques, viral loads below the detectable limit of the assay, or diminished upper airway shedding of the virus ^22-25^. Finally, the role of specific antibodies in terms of protection against reinfection and persistence over time is currently not adequately defined.

In the beginning of the pandemic, lateral flow tests were primarily employed to perform serological SARS-CoV2 testing ^26^. However, some of these tests have been criticized for poor sensitivity and specificity ^27^. Poor specificity has been suspected to occur due to cross-reactivity with the antibody response to endemic coronaviruses causing the common cold (i.e., HCoV-229E, -NL63, -OC43 and - HKU1) ^28^. Some of these cross-reacting antibodies, however, have actually shown neutralizing activity against SARS-CoV-2 ^29^. Since the antibody response in SARS-CoV-2 infection needs 2 to 4 weeks to develop, false negative antibody tests can occur due to insufficient duration between the onset of clinical symptoms and the time of blood sampling or insensitive measurement techniques that require large quantities of antibodies for a positive result ^6,12^. At the moment, it is not known how long the antibody titers persist. A recent Cochrane review stated that there are currently not enough studies available for antibody tests done after 35 days of symptom onset ^30^.

Whereas ELISA tests were available relatively early during the pandemic, the supply of these tests has been relatively scarce ^31,32^. Only recently have assays using chemiluminescence (CLIA) become available, with only a few validation data published so far ^33,34^. These assays are available in large quantities and can be analyzed in high-throughput settings. The different assays are directed against different specific antigens, i.e., the internal nucleocapsid (N) antigen or the surface spike protein (S1 or S1/S2) ^27^. Not all SARS-CoV2 antibodies exhibit neutralizing properties. However, it has been shown that antibodies against nucleocapsid antigen and the receptor-binding domain (RBD) of the spike protein have a high correlation to virus neutralization titer ^7^.

The predictive values of negative and positive results depend on the pretest probability of the presence of COVID-19 disease. In low-prevalence settings (<5% seropositivity), positive predictive values are critically dependent on specificity ^35^. Since the specificity of SARS-CoV2 antibody tests has been the subject of debate, false positive results can occur in low-prevalence settings ^36^. False positive assay results are particularly problematic in low prevalence settings, as these could lead concerned individuals into a false sense of security and increase their risk of contracting COVID-19 disease through unsafe behavior ^13^.

We aimed to investigate the diagnostic specificity and sensitivity of newly released chemiluminescence immunoassays (CLIAs). We investigated these assay formats in a cohort of confirmed COVID-19 patients to assess diagnostic sensitivity. Subsequently, we assessed the diagnostic specificity of these tests in a cohort of healthcare workers and a cohort of healthy blood donors. Finally, we assessed the analytical specificity of the different assays in samples collected prior to the COVID-19 pandemic, which may potentially contain cross-reacting antibodies. In a model, we finally related the identified diagnostic characteristics to negative and positive predictive values depending on the pretest probability of having had COVID-19 in the personal history.

## Methods

### Study setting and study population

This is a study on diagnostic tests used for detection of SARS-CoV-2 antibodies in individuals originating from Switzerland and Liechtenstein. Anonymized samples originating from four different types of patients were investigated with three CLIAs. The first group of patients was retrospectively assembled from Liechtenstein and Swiss patients having their laboratory evaluations sent to labormedizinisches zentrum Dr. Risch in Vaduz (Liechtenstein) and Buchs (Switzerland) and consisted of COVID-19 patients (mainly outpatients) whose serum was drawn after COVID-19 disease was confirmed by RT-PCR between March 2^nd^ and April 23^rd^, 2020 (n=145). This group was used for determination of sensitivity. The method of RT-PCR measurement has been conducted as reported elsewhere ^37^. The patients from Liechtenstein were prospectively and consecutively enrolled within a national COVID-19 cohort, whereas the Swiss patients consisted from a retrospectively assembled convenience sample, both detailed in references ^37,38^, The second group of individuals was prospectively assembled and consisted of consecutive healthy blood donors from the Blutspendedienst Graubünden without clinical suspicion of COVID-19 providing blood for testing SARS-CoV2 antibodies from April 15^th^ to May 4^th^, 2020 (n=191). In addition to fulfilling the normal criteria of blood donation, these blood donors specifically responded they had had no had flu-like symptoms or contact with a known COVID-19 patient during the past 14 days. The third cohort was prospectively assembled within a study setting from the Kantonsspital St. Gallen and consisted of healthcare workers providing a blood sample for detecting SARS-CoV2 antibodies between March 19^th^ and April 3^rd^, 2020 (n=1002) as described elsewhere ^39^. Hospital admissions in this region peaked in the second week of April 2020. The blood donors and the healthcare workers were used to determine specificity. Since these 2 groups did not have RT-PCR testing available, samples with at least two of the three chemiluminescence assays positive were excluded from this analysis, assuming that these individuals had occult SARS-CoV-2 infection. Such an orthogonal testing approach for SARS-CoV-2 antibodies has been reported to have a very high positive predictive value for COVID-19 and therefore is suited to reliably exclude individuals with recent COVID-19 infection ^40-42^. Characteristics of the excluded 10 seropositive individuals among the healthcare workers are detailed elsewhere (i.e. 6 individuals had all three assays positive, 3 had CMIA and ECLIA positive, one had CMIA and LIA positive) ^39^, whereas 4 individuals were excluded among the blood donors (one individual with positive SARS-CoV-2 RT-PCR result 24 days before sampling, three asymptomatic individuals). In the excluded blood donors, all three chemiluminescence assays were positive. The fourth group of samples consisted of historical samples sent to the labormedizinisches zentrum Dr. Risch in Buchs (Switzerland) that were known to have an active or reactivated specific viral disease (Epstein–Barr virus, EBV, n=9; cytomegalovirus, CMV, m=7; other common-cold coronaviruses: HKU1, NL63, OC43, 229E, n=12) to explore any cross-reactivity causing false positive results in SARS-CoV2 serology. Endemic coronavirus disease was diagnosed during 2019 in 10 cases, in January 2020 in 1 case, and in mid-February 2020 1 case, 8 days before the first case of COVID-19 was reported in Switzerland. The last serum sample of patients with endemic coronavirus disease was collected on March 2^nd^, 2020, which was 7 days after the first case in Switzerland was identified. Samples from patients with active EBV (VCA IgM positive, EBNA IgG negative) as well as active or reactivated CMV infection (IgG positive, IgM positive) were all drawn in 2019, i.e., before COVID-19 was first diagnosed in Switzerland and Liechtenstein. The study protocol was verified by the cantonal ethics boards of Zurich (BASEC Req-20-00587) and Eastern Switzerland (EKOS; BASEC Nr. Req’s 2020-00502 and 2020-00586). Whereas the cohort with healthcare workers provided written informed consent, informed consent for performing laboratory analysis on anonymized samples in the other three groups was waived. The study was taking into account the STARD guidelines ^43^.

### Data collection and measurements

For each serum sample, the age and sex of the individual, the type of clinical setting, and the number of days after first positive RT-PCR it was drawn (if applicable) were available. The sera employed for testing were either fresh or stored at −25°C for less than 18 months. The antibodies were tested on the following diagnostic platforms: COBAS 6000 (Roche Diagnostics, Rotkreuz, Switzerland), Abbott Architect i2000 (Abbott Diagnostics Baar, Switzerland), and Liaison XL (Diasorin, Luzern, Switzerland). The Roche Diagnostics assay (Elecsys^®^ Anti-SARS-CoV-2; ECLIA) employs a recombinantly engineered nucleocapsid antigen for detection of total immunoglobulin. The molecular target from the Abbott Diagnostics assay (SARS-CoV-2 IgG; CMIA) is also the nucleocapsid antigen, and it measures specific IgG levels. The Diasorin assay (LIAISON^®^ SARS-CoV-2 S1/S2 IgG; LIA) measures specific IgG directed against S1/S2 antigens. To further elucidate the effects of any cross-reacting antibodies in the 3 chemiluminescence assays in the samples with EBV, CMV, or following endemic coronavirus, we also employed the Euroimmun SARS-CoV-2 ELISA (Euroimmun, Luzern, Switzerland) to measure specific IgG and IgA on a DSX instrument (Dynex Technologies, Denkendorf, Germany). The coefficients of variation (CV’s) of the employed methods in our hands were 2.7% for ECLIA, 3.6% for CMIA, 5.4% for LIA, 4.6% for IgG ELISA, and 3.6% for IgA ELISA.

### Statistical methods

Specificity was determined in the samples originating from blood donors and healthcare workers. The sensitivity of the different assays was assessed in the group of patients with RT-PCR confirmed COVID-19 disease. The following cut-offs provided by the manufacturers were employed: a COI (COI= cutoff index) >0.9 for ECLIA, a S/C value (S/C=extinction of the patient sample divided by the extinction of the calibrator) >12 for LIA, a S/C value ≥1.4 for CMIA, and a S/C value of ≥1.1 in the ELISA. We also applied alternative cut-offs (half and double of the manufacturers’ cut-offs), in order to better understand the relationship between signal strength and associated diagnostic characteristics. Positive and negative likelihood ratios (LR) were calculated for the different cut-off levels ^44^. Regarding interpretation of LR, it is generally acknowledged that a +LR >10 as well as a –LR <0.1 generate large and often conclusive changes from the pretest to the posttest probability. A +LR between 5 and 10 as well as a –LR between 0.1 and 0.2 generate moderate shifts from the pretest to the posttest probability, whereas a +LR between 2 to 5 and a –LR from 0.2 to 0.5 generate small but sometimes important changes from the pretest to the posttest probability ^44^. Receiver operating characteristic (ROC) curves with the area under the curves (AUCs) calculated as an indicator of diagnostic accuracy. The AUCs for the different parameters were compared by the method of Hanley and McNeil. Positive and negative predictive values for each of the employed assays were then plotted as a function of pretest probability, as described earlier ^45^. Continuous variables are given as medians and interquartile ranges [IQRs], whereas proportions are given as percentages together with 95% confidence intervals (CIs). Agreement between the two methods was assessed by Cohen’s kappa. The associations between variables were calculated with Spearman rank correlation. P-values <0.05 were considered statistically significant. Medcalc version 18.11.3 (Mariakerke, Belgium) and Microsoft Excel 2016 MSO (16.0.8431.2046) (Microsoft Inc, Seattle, USA) were used for statistical and graphical computations.

## Results

### Baseline characteristics

The group of RT-PCR confirmed COVID-19 patients consisted of 145 individuals with a median age of 46 years (IQR [30,58] years), and 79 of the patients, i.e., 48% (95% CI [40,56]), were female. The sera were taken after a median of 47 IQR [40,54; minimum 21, maximum 66] days after first presentation with suspected COVID-19. The blood donors had a median age of 44 years IQR [28,53], and 90 of the 191 donors, i.e., 47% (95% CI [40,54]) were female. In the cohort of healthcare workers, the median age was 38 years (IQR [30,49]), and 753 (75.1%, 95% CI [72.4,77.7]) were female. The cohort with prior EBV, CMV or endemic coronavirus had a median age of 31 years (IQR [16,60]), and 17/30 (57%, 95% CI [39,73]) were women. Serum samples of patients with common cold caused by endemic coronavirus had their samples taken a median of 94 (IQR [30,235]) days after diagnosis. As shown in Figure 1 a-c, antibody titers were different between COVID-19 patients and individuals without COVID-19. The correlation between the 3 different assays was highly significant (p<0.0001). The agreement of the methods when we used the manufacturers’ cut-offs showed a kappa value of 0.92 between CMIA and LIA, 0.96 between CMIA and ECLIA, and 0.94 between LIA and ECLIA. In the COVID-19 patients, there was a significant inverse association between days after RT-PCR and S/C in CMIA (r=-0.25, p=0.003), but not in LIA or ECLIA.

**Figure 1.**
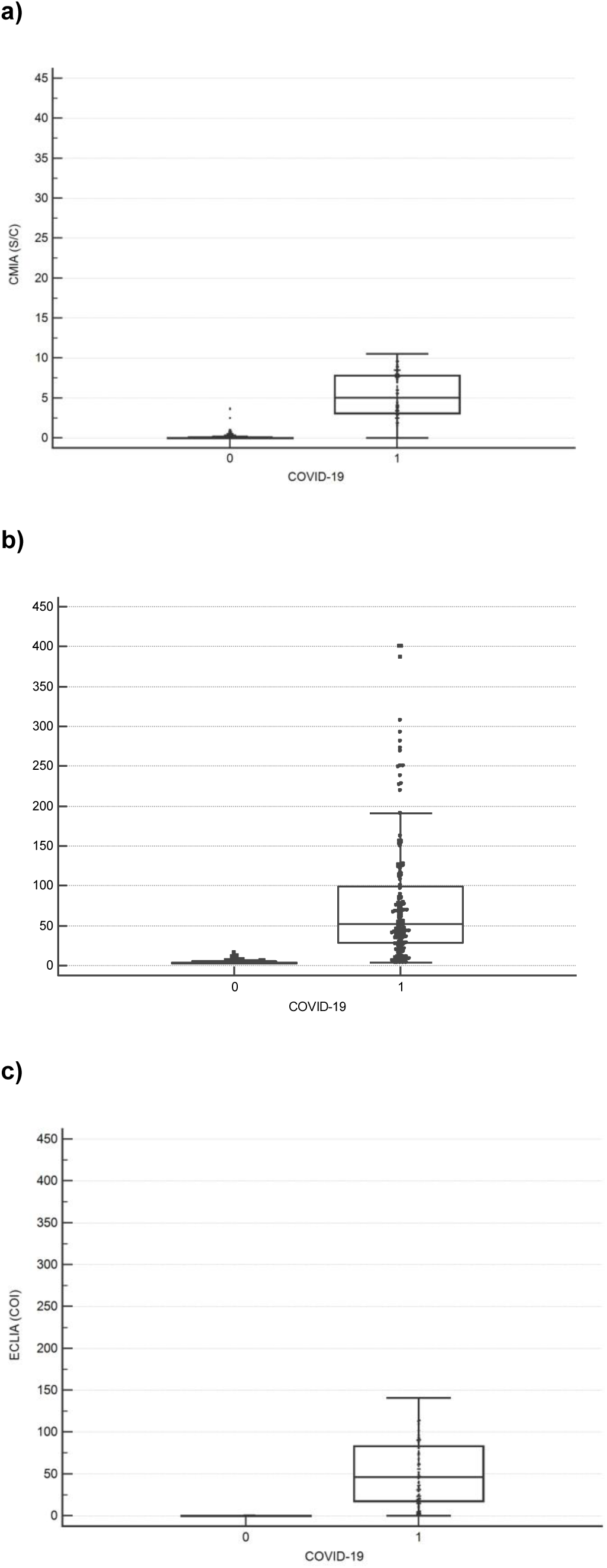
SARS-CoV-2 antibody titers of three different chemiluminescence assays in individuals with and without COVID-19: a) CMIA, b) LIA, c) ECLIA. COVID-19=0: individuals without evidence of COVID-19; COVID-19=1: patients with RT-PCR confirmed COVID-19;

### Diagnostic specificity and sensitivity at the manufacturers’ cut-offs

When looking at specificities of the different assays at the cut-offs provided by the manufacturers within the group of blood donors and healthcare workers without COVID-19, the following characteristics were observed: 99.5% specificity (95% CI [98.9,99.8], i.e., 1187/1193 individuals; 3 false positives from blood donors, 3 false positives from healthcare workers) for the CMIA, 99.7% specificity (95% CI [99.3,99,9], i.e., 1190/1193 individuals; 1 false positive from blood donors, 2 false positives from healthcare workers) for the LIA, and 99,91% specificity (95% CI [99.5,99.98], i.e. 1192/1193 individuals; 1 false positive from healthcare workers) for the ECLIA. There was no overlap between the participants with false positive results across the three assays. The respective sensitivities were 93% (95% CI [88,96], i.e., 135/145) for the CMIA, 90% (95% CI [84,94], i.e., 130/145) for the LIA, and 96% (95% CI [91,98], i.e., 139/145) for the ECLIA. When performing ROC analysis on all COVID-19 cases and healthy controls, the AUCs of the different assays in detecting COVID-19 disease were 0.984 (95% CI [0.976,0.99]) for CMIA and 0.982 (95% CI [0.974,0.989]) for both the LIA and the ECLIA (curves not shown). There were no significant differences between the AUCs of the three assays.

### Multilevel likelihood ratios

We then calculated multilevel likelihood ratios at the manufacturers’ cut-off levels as well as at half and double the manufacturers’ cut-off levels, i.e., a S/C of 0.7, 1.4 and 2.8 for the CMIA, respectively, a S/C of 6, 12, and 24 for the LIA, and a COI of 0.5, 1, and 2 for the ECLIA. Table 1 illustrates the different positive (+LR) and negative likelihood ratios (-LR). Regarding diagnostic value of the LR, the investigated serological tests can be considered to offer meaningful diagnostic characteristics. Of note, clinical characteristics are already meaningful at lower cut-off levels than recommended by the manufacturer.

**Table 1.** Multilevel likelihood ratios as well as sensitivity and specificity are given for the different tests at different cut-off levels (manufacturers’ cut-offs, half and double the manufacturers’ cut-offs) for ruling in or ruling out a COVID-19 diagnosis. +LR= positive likelihood ratio; -LR= negative likelihood ratio. CMIA= chemiluminescence microparticle immunoassay; LIA= luminescence immunoassay; ECLIA=electrochemiluminescence immunoassay; Sens= sensitivity; Spec= specificity, CI= confidence interval.

|  | **+LR at**<br>**cut-off/2**<br>*(Spec, [95% CI])* | **-LR at**<br>**cut-off/2**<br>*(Sens, [95% CI])* | **+LR at**<br>**cut-off**<br>*(Spec, [95% CI])* | **-LR at**<br>**cut-off**<br>*(Sens, [95% CI])* | **+LR at**<br>**2x cut-off**<br>*(Spec, 95% CI)* | **-LR at**<br>**2x cut-off**<br>*(Sens, 95% CI)* |
| --- | --- | --- | --- | --- | --- | --- |
| **CMIA** | 60<br>*(98.4 [97.5,99.0])* | 0.043<br>*(95.8 [91.0-98.4])* | 374<br>*(99.8 [99.3,99.9])* | 0.056<br>*(94.4 [89.2,97.5])* | 485<br>*(99.8 [99.4,100])* | 0.18<br>*(81.7 [74.3,87.7])* |
| **LIA** | 82<br>*(98.8 [98.0,99.4])* | 0.035<br>*(96.5 [92.1,98.9])* | 351<br>*(99.8 [99.3 - 99.9])* | 0.12<br>*(88,2 [81.8,93.0])* | 1027<br>*(100 [99.7,100])* | 0.14<br>*(86.1 [79.4 - 91.3])* |
| **ECLIA** | 575<br>*(99.8 [99.4,100])* | 0.035<br>*(96.5 [92.1,98.9])* | 958<br>*(99.9 [99.5,100])* | 0.042<br>*(95.8 [91.2,98.5])* | >958<br>*(100 [99.7,100])* | 0.042<br>*(95.8 [91.2,98.5])* |

### Operational test characteristics

The positive predictive values (PPVs) for three different cut-offs (manufacturers’ cut-offs and half as well as double the manufacturers’ cut-off values are shown in Figures 2a (CMIA), 2b (LIA), and 2c (ECLIA)). It can be seen that ECLIA, especially in low pretest probability settings, due to the high specificity has somewhat higher PPVs than the other assays: at the manufacturer’s cut-off, the PPV was 97% at a pretest probability of 3 percent, whereas at half of the manufacturer’s cut-off, the PPV at a pretest probability of 3% was 95%. For the LIA, the PPV at a pretest probability of 5% was 95%, whereas the same PPV at half of the manufacturer’s cut-off was achieved with a pretest probability of 18%. At double the manufacturer’s cut-off, a PPV of 95% in the LIA was achieved with a pretest probability of 2%. Finally, with the CMIA, at the manufacturer’s cut-off, a PPV of 95% was achieved at a pretest probability of 5%, whereas at half of the manufacturer’s cut-off, the same PPV was seen at a pretest probability of 23%. At double the manufacturer’s cut-off, the CMIA had a PPV of 95% at a pretest probability of 4%.

**Figure 2.**
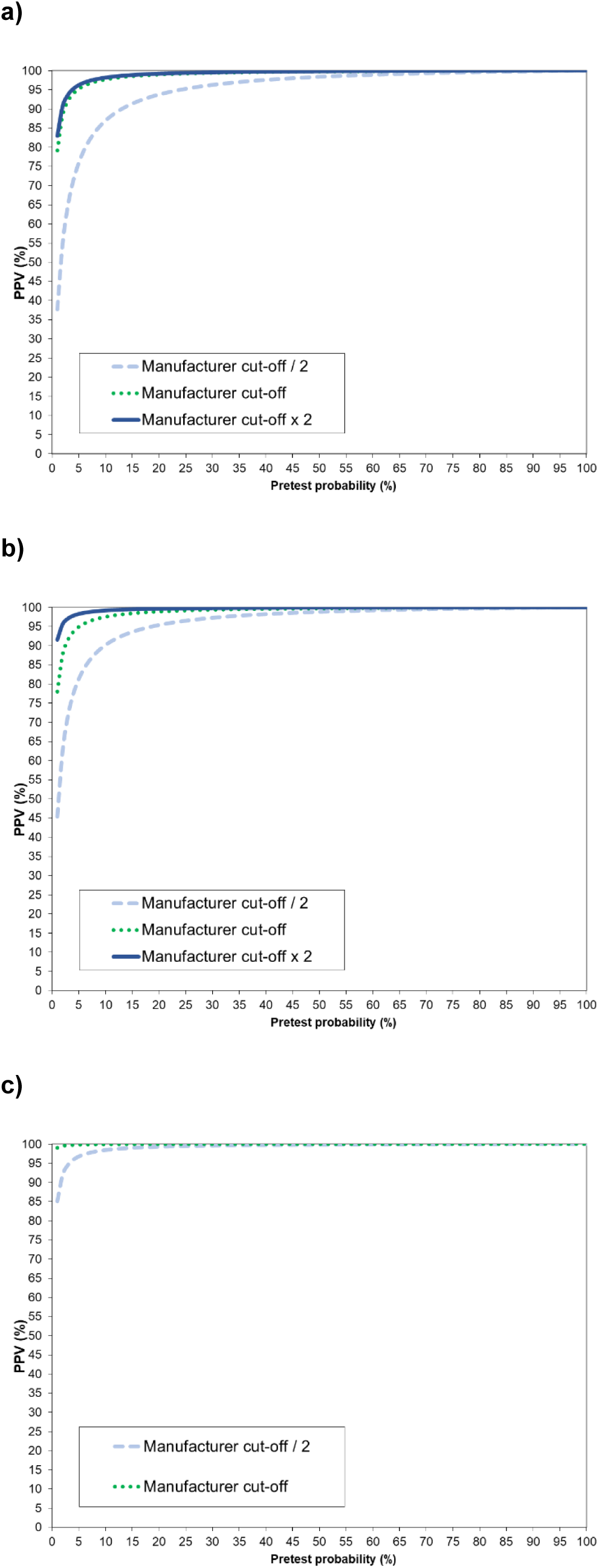
Positive predictive values (PPVs) of SARS-CoV-2 antibody titers at three different cut-offs (manufacturers’ cut-offs, half and double the manufacturers’ cut-offs) over the whole range of possible pretest probabilities. Three different chemiluminescence assays were assessed: a) CMIA, b) LIA, c) ECLIA

**Figure 3.**
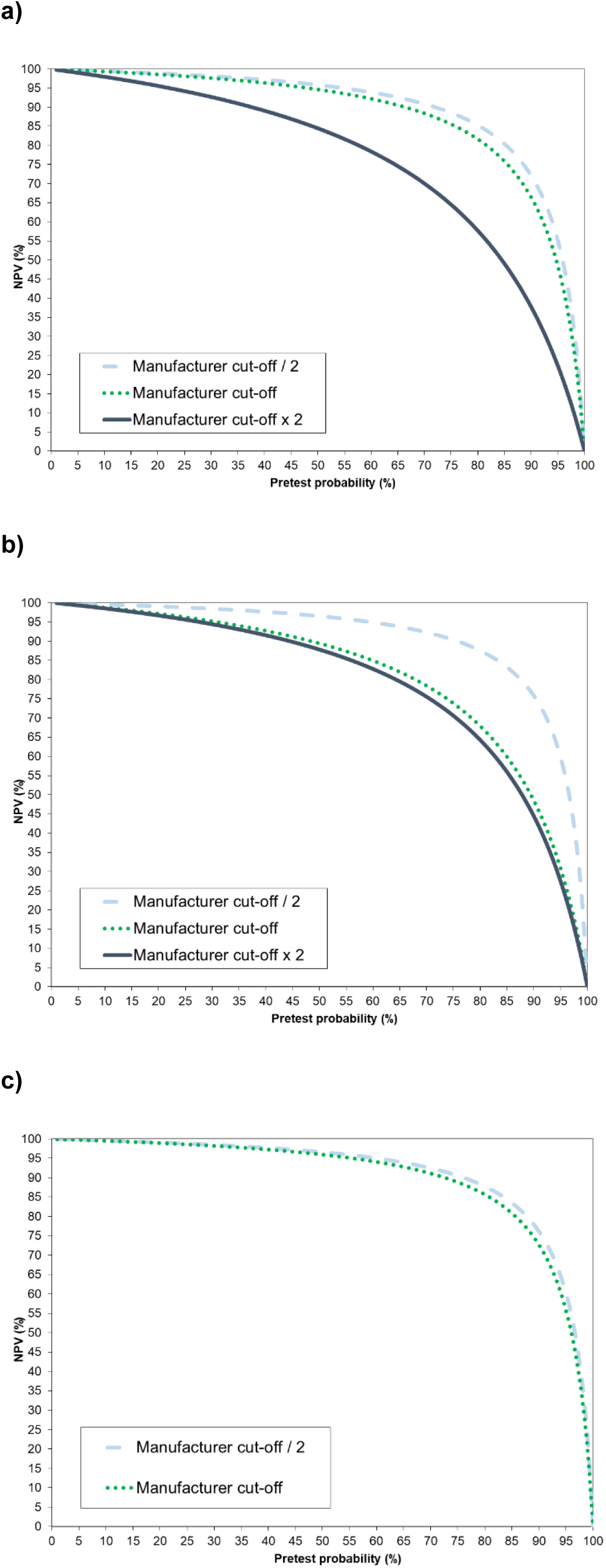
Negative predictive values (NPVs) of SARS-CoV-2 antibody titers at three different cut-offs (manufacturers’ cut-offs, half and double the manufacturers’ cut-offs) over the whole range of possible pretest probabilities. Three different chemiluminescence assays were assessed: a) CMIA, b) LIA, c) ECLIA.

Regarding negative predictive values (NPVs), the ECLIA due to the highest sensitivity had a NPV of 99% up to a pretest probability of 26% at the manufacturer’s cut-off. At half of the manufacturer’s cut-off, a NPV of 99% was seen up to a pretest probability of 30%. With the LIA, the NPV at the manufacturers’ cut-off was at 99% up to a pretest probability of 11%, whereas at half of the manufacturer’s cut-off, the NPV was at 99% up to a pretest probability of 30%. In the CMIA, the NPV at the manufacturer’s cut-off was 99% up to a pretest probability of 21%, whereas at half of the cut-off, the NPV was 99% up to a pretest probability of 26%.

### Analytical specificity

None of the assays showed positive antibodies in sera from patients with active EBV or CMV disease. Of the sera taken after endemic coronavirus infection, 4 were infected with RC229E, 3 were infected with RCNL63, 2 were infected with RCHKU1, 2 had an infection with RCOC43, and one patient had both RC229E and RCNL63. None of the patients showed antibody positivity in the ECLIA. In three patients following infection with different coronaviruses, the CMIA showed an S/C result of 0.4 (infection with RCHKU1; SARS-CoV-2 ELISA IgG 0.1; IgA 0.3), 0.5 (infection with RCOC43; SARS-CoV-2 ELISA IgG 0.1; IgA 0.1), and 0.1 (infection with RC229E; SARS-CoV-2 ELISA IgG 0.7; IgA 4.1), whereas the other samples did not reveal a detectable signal. The sample of a patient with RCOC43 infection with a detectable CMIA signal also had a detectable LIA signal of 6.1, whereas all other samples had unmeasurable signals. All together, even if SARS-CoV-2 antibody titers following endemic coronavirus infections were below the manufacturers’ cut-offs for positivity, 3 samples exhibited detectable antibodies in the CMIA assay, one of which was also in the LIA assay.

## Discussion

The present study investigated the analytical (analytical specificity), diagnostic (diagnostic sensitivity and specificity), and operational characteristics (likelihood ratios, predictive values) of three highly automated assays for the detection of antibodies against SARS-CoV-2. These characteristics were investigated after a median of 47 days after PCR-positivity. Although the three test formats showed comparable diagnostic accuracy, test performance varied regarding positive predictive values in the low-prevalence setting. Finally, the investigated tests did not show interference with two commonly encountered infections, i.e., EBV and CMV. There was, however, cross-reactivity in some patients with proven previous infection by endemic coronaviruses for two of the three analyzed tests.

The timing of antibody development in COVID-19 disease is crucial. Long and colleagues demonstrated that IgG or IgM can be detectable in approximately 60% of patients at days 5-7 after symptom onset, in 95% of patients after 12-14 days and in 100% of patients after 17 to 19 days ^6^. Tang and colleagues demonstrated a seroconversion rate of 93.8% for CMIA and 89.4% for ECLIA 14 days after symptom onset and later ^46,47^. A systematic review performed by the Cochrane collaboration regarding the usefulness of antibody tests to diagnose current or past infection with SARS-CoV-2 stated that there are too few studies available to estimate test sensitivity beyond 35 days after symptom onset ^30^. The present study, with a median of 47 days after presentation for symptomatic COVID-19 disease, therefore fills an important gap.

Laboratory results are interpreted by combining pretest probabilities with diagnostic characteristics to obtain negative and positive predictive values according to Bayes’ theorem ^48^. To the best of our knowledge, there is no clinical score available to predict the pretest probability of COVID-19 based only on clinical symptoms and history without any laboratory or imaging results ^49^. Such a score based on clinical history taking would improve the interpretation of COVID-19 serology results, not least in a retrospective setting, where accurate laboratory results are not available because no blood has been drawn. However, there are several other proxies to assess pretest probabilities. In the Swiss and Liechtenstein setting, several situations can be described, in which the risk for patients to contract COVID-19 can be quantified ^37^: 32% for household contacts, 13% of close working contacts, and 12-25% of patients with fever >38°C and respiratory symptoms. Knowing the temporal and regional seroprevalence of the region also allows for an approximation of pretest probability even without knowing the clinical symptoms of a specific patient ^50^.

The predictive values depend not only on the pretest probability but also on the sensitivity and specificity of a test at a given cut-off. Negative results using the manufacturers’ cut-offs had an NPV of 99% up to a pretest probability of 11% to 26%. At half of the manufacturer’s cut-off, past COVID-19 could be ruled out with a probability of 99% up to a pretest probability of 26% (CMIA) to 30% (LIA, ECLIA). These pretest probabilities are in the range of those observed in symptomatic individuals. Thus, serology done in patients with clinical symptoms three or more weeks before drawing blood together with manufacturers’ and modified manufacturers’ cut-offs can be considered safe to exclude COVID-19 disease. Nevertheless, serological non-responders can occur with a frequency of approximately 3% and may go overlooked. False negative results, if they occur at a low frequency, can be considered of minor importance because they are not expected to change an individual’s behavior in response to its potential damage.

Regarding PPV, which is the probability that a positive result indicates a past infection with SARS-CoV-2, the ECLIA with the manufacturer cut-off provided the best operational characteristics at low pretest probabilities ≤ 5%. In this assay, the PPV at 3% pretest probability was 97%, which raises the question whether positive ECLIA results should be confirmed by an orthogonal testing approach ^40,51,52^. When taking the lower limit of the 95% CI of the specificity, i.e. 99.5%, the PPV at 1% pretest probability is 66%, and 91% at 5% pretest probability. We therefore recommend continuing with the confirmation of positive ECLIA results with a second assay ^40^. Our data also show that at the manufacturer cut-offs, the CMIA and LIA need confirmatory testing in low pretest probability (<5%) settings. Otherwise, the risk of false positive results is larger than 5%. The problem of false positive results is that they can change the behavior of individuals in that they will not adhere to hygienic and distancing measures after the (false) positive result has been disclosed. This puts individuals at increased risk for contracting COVID-19, with its potentially devastating consequences ^13^. The medical laboratory thus has a great responsibility to deliver meaningful results to the patients and their physicians.

Common recommendations on likelihood ratios state that +LR>10 and -LR<0.1 generate meaningful changes from pretest to posttest probability ^44^. Our findings suggest that all three assays provide meaningful information, even at half of the manufacturers’ cut-off values. The figures of pretest probabilities against NPV illustrate that taking half of the manufacturer cut-off as a decision limit will still safely rule out COVID-19: a cut-off as low as half of the manufacturer cut-off will correctly detect 98.9% (95% CI [98.1,99.4]) (13/1193) of the COVID-19-negative cases in the CMIA. The respective proportions are similar for the LIA (98.8%, 95% CI [98,99.3], (14/1193) and ECLIA (99.7%, 95% CI [99.3,99.9], 3/1193). For ruling out COVID-19, modified decision limits thus seem appropriate.

Looking at the relationship between pretest probability and PPV at a cut-off half of the manufacturer’s cut-off illustrates that such a cut-off does not reliably diagnose past COVID-19 in low test probability settings. However, using an orthogonal testing approach recommended by the U.S. Food and Drug Administration ^41^, i.e., an ECLIA result of 0.6 together with an LIA result of 6.1, would lead to a PPV of 99.1% at a pretest probability of 1%. A similar result would be achieved with a CMIA result of 0.7 together with an LIA result of 6.1, which at a pretest probability of 2% results in a PPV of 99%. This example illustrates that the introduction of gray zones should be considered not only for ELISA but also for chemiluminescence immunoassays when offering SARS-CoV-2 testing to clinicians and public health professionals ^53^.

The fact that we found some cross-reactivity in patients with prior endemic coronaviruses raises the question whether this finding represents analytical cross-reactivity or reflects cross-reactive immunity conferred by recent endemic coronavirus disease. We did not conduct neutralization assays to clarify this issue. However, Ng and colleagues reported that patient sera from human coronaviruses variably reacted with SARS-CoV-2 S-antigen and nucleocapsid antigen, but not with the S1 subunit ^29^. These patient sera exhibited neutralizing activity against SARS-CoV-2 S pseudotypes according to the levels of SARS-CoV-2 S-binding IgG and with efficiencies comparable to those of COVID-19 patient sera ^29^. In our study, we had one case with a LIA signal (targeting S1-S2 antigen) and a CMIA signal (targeting nucleocapsid antigen) without a signal in the IgG ELISA (which is targeted against S1). This patient had his serum taken in September 2019 after infection with RCOC43 was diagnosed in January 2019. We are convinced that identifying patients with endemic coronaviruses with potential neutralizing cross-reactivity will be of increasing importance in the future. Whereas CMIA and LIA could play a role in identifying such individuals, ECLIA is not expected to help further in this aspect, as none of the patients with endemic coronavirus had a measurable antibody response. The measurements in the healthcare workers as well as in the blood donors suggest that such constellations do not occur often: 1147/1193 (i.e., 96.1 (95% CI [94.9,97.1])) have unmeasurable antibody titers in the LIA, whereas this frequency amounts to 921/1193 (i.e., 77.2% (95% CI [74.7,79.5])) in the CMIA.

Our study has strengths and limitations. A strength is that specificity has been assessed in a large group of 1193 individuals without evidence of COVID-19. Such an approach offers the possibility to describe the specificity with relatively narrow confidence intervals. A further strength is that we investigate several potential cut-offs for clinical decision making. A limitation of the study is that samples employed for evaluation of specificity were selected from contemporary and not pre-pandemic participants. Two positive serology results in an orthogonal testing approach cannot provide 100% certainty that any remaining false positives are truly false positive. Nevertheless, we demonstrate that combination of two positive results with chemiluminescence assays has a very high positive predictive value even at low pretest probabilities, comparable to that known from molecular methods ^42^. Further, inclusion of the index serology tests as part of the reference standard definition of absence of disease carries a risk of bias in results. At low pretest probabilities, such a bias might have a considerable impact on positive predictive values. There are several factors mitigating such a misinterpretation: a.) utilizing an orthogonal testing approach in initially positive results, b.) the fact that even in the first wave of COVID-19, COVID-19 prevalence in some regions of Europe was already more than 10%, which cannot be considered low ^50^. The specificities identified in our study from contemporary samples are comparable to those obtained in a similar study on pre-pandemic samples ^54^. The study is finally limited by the fact that we did not have a full clinical description of the COVID-19 patients included in this study, and patients hospitalized due to COVID-19 represented a minority of the COVID-19 patients. However, since the majority of patients subjected to serology will originate from the outpatient setting, we believe that this fact actually strengthens our findings. In sum, we think that the limitations do not invalidate our findings.

In conclusion, we evaluated the serology in patients at a median of 47 days following the first presentation of suspected COVID-19 infection and selected individuals without COVID-19 during the pandemic. We found that the diagnostic accuracy of the three investigated assays was comparable. Assay cut-offs have not been designed for orthogonal testing. Introducing gray zones at half of the manufacturers’ cut-offs is suggested. These differentiated cut-offs would allow for safer ruling out or ruling in of past COVID-19 infections. Such an approach would allow us to more appropriately select samples for further testing in an independent assay in an orthogonal testing algorithm. We propose that our findings be replicated in other populations.

## Data Availability

The data used to support the findings in this study will be available from the corresponding author upon request.

## Acknowledgments

The help of Toni Schönenberger and Walter Frehner in identifying samples is acknowledged.

## CRediT author statement

**Myriam Weber:** Conceptualization, Methodology, Formal analysis, Writing - Original Draft; **Martin Risch:** Conceptualization, Methodology, Formal analysis, Writing - Original Draft; **Sarah Thiel:** Data curation, Validation, Resources; **Kirsten Grossmann:** Resources; **Susanne Nigg:** Resources; **Kirsten Nadja Wohlwend**: Investigation, Validation, Resources; **Thomas Lung:** Validation, Resources; **Dorothea Hillmann:** Investigation, Validation, Project administration, Resources; **Michael Ritzler:** Resources; **Francesca Ferrara:** Resources; **Susanna Bigler:** Validation, Resources; **Konrad Egli:** Validation, Resources; **Thomas Bodmer:** Validation, Resources; **Mauro Imperiali:** Validation, Resources; **Yacir Salimi:** Validation, Resources; **Felix Fleisch:** Resources; **Alexia Cusini:** Resources; **Sonja Heer:** Resources; **Harald Renz:** Conceptualization; **Matthias Paprotny:** Supervision, Resources, Writing - Original Draft; **Philipp Kohler:** Validation, Writing - Original Draft; **Pietro Vernazza:** Conceptualization, Funding acquisition, Writing - Original Draft; **Lorenz Risch:** Conceptualization, Methodology, Funding acquisition, Supervision, Resources, Writing - Original Draft; **Christian R. Kahlert:** Conceptualization, Methodology Supervision, Resources, Writing - Original Draft; **All authors:** Writing-Reviewing and Editing

